# Cross-reaction of sera from COVID-19 patients with SARS-CoV assays

**DOI:** 10.1101/2020.03.17.20034454

**Authors:** Wei Yee Wan, Eng Hong Seng, Siew Hoon Lim

## Abstract

**Background:** The SARS-CoV-2 shares 74.5% genome identity with SARS-CoV, both exhibiting a similar well conserved structure. Therefore, antibodies produced in COVID-19 and SARS patients should not be that dissimilar. We evaluated SARS-CoV test assays to detect for the presence of antibodies to SARS-CoV-2 and tried to determine the timing of appearance of these antibodies by testing serial sera from these patients.

**Methods:** Tests were carried out using ELISA (total antibodies) and indirect immunofluorescence (IIFA) (IgM & IgG) methods on serial sera from patients confirmed with SARS-CoV-2 infection.

**Results:** Cross-reactivity was seen in these two test assays with sera from COVID-19 patients and was detected in 6 out of 7 patients from 7 days after onset of symptoms. Five of the patients had detectable antibodies by the 3^rd^ week into their illness and there was evidence of seroconversion in 4 patients. The IIFA method was marginally more sensitive compared to the ELISA assay, however the IIFA IgM test was not useful in the early phase of the illness with poor sensitivity.

**Conclusions:** Existing diagnostic assays for SARS-CoV can detect antibodies in patients who were diagnosed with COVID-19. These assays maybe be utilized as an interim measure in epidemiological investigations for contact tracing and to determine the extent of community spread of this new emerging virus pending the availability of specific serology tests for SARS-CoV-2.

## Introduction

SARS-CoV-2 is a new zoonotic coronavirus (CoV) that emerged in Wuhan, China which was first reported on the 31^st^ December 2019. As the number of cases of COVID-19 increases, there is an urgent need to understand this outbreak that looks set to spread to several countries around the world. The total number of cases is probably a gross underestimate, as patients may present with the symptoms of the common cold, and therefore remain undiagnosed due to the mild nature of this illness in the majority. Poorer resourced countries may also not have the capability to equip themselves with complex molecular diagnostic setups and thus outbreaks in those areas may go undetected for some time and the number of cases under reported.

The SARS-CoV-2 is a SARS-related virus with 74.5% genome identity to SARS-CoV.^1^ The similarities between these two viruses were described comprehensively in a recent published article by Xu et al.^2^ For structural proteins, including the nucleocapsid (N), matrix (M), and envelope (E), high within-group conservation was maintained, with more modest similarity seen across the entire CoV family. In contrast, the accessory proteins that distinguish CoV infections from each other with high variability across the family, allow viruses to adopt to current and novel hosts.^3^ In a study which described the difference in amino acid substitutions of different proteins for SARS-CoV-2 compared to SARS-CoV, it was found that there were no substitutions that occurred in nonstructural protein 7 (nsp7), nsp13, envelop, matrix and accessory proteins p6 and 8b.^4^ The N protein for SARS-CoV-2 has ∼90% similar amino acid identity to the SAR-CoV N protein and hence the SARS-CoV-2 antibodies against the N protein would likely recognize and bind the SARS-CoV N protein as well.^5^ Furthermore, a study by Zhou et al, showed that the SARS-CoV-2 could be cross-neutralized by horse anti-SARS-CoV serum at dilution 1:80, confirming the relationship of the 2 viruses.^6^

Based on this knowledge, we postulated that the antibodies produced by COVID-19 patients should result in cross-reactivity to the SARS-CoV total antibody ELISA & IIFA tests which utilizes whole SARS-CoV infected cells as the antigen substrate.

Patient’s consent and ethical approval from the Ethics Committee were not required as per the CIRB Research committee’s guidelines and advice for the evaluation of this assay which used existing anonymized human biological materials for test validation purposes and there is no prospective collection of clinical, pathological and demographic information.

## Methods

We identified SARS-CoV-2 positive cases which were confirmed by molecular testing of respiratory specimens by real-time RT-PCR according to the published protocol by Corman et al.^7^ We retrieved residual samples left over from biochemical tests to obtain serial sera for these patients. The Biochemistry department removed all patient identifiers and assigned random numbers to each patient, and also included the number of days after onset of illness for each of the retrieved specimens based on the information obtained from the Infectious Diseases team. For negative controls, 10 samples which were sent for unrelated virology tests from 2 different groups of patients were randomly selected. The first group consists of 5 sera collected 5 years ago from our archive and the other group, another 5 sera from patients who were tested negative on two occasions for SARS-CoV-2 as part of the enhanced surveillance for patients who presented with pneumonia but did not fulfil the criteria of suspected SARS-CoV-2 infection during this outbreak period.

We performed two serological test methods on the selected samples using the SARS-CoV total antibody ELISA test as described by Ksiazek et al^8^ and Anti-SARS CoV Indirect Immunofluorescence test (IIFT) (IgM & IgG) by Euroimmun (Germany) according to the specified protocols and manufacturer’s instructions respectively. Both these tests had previously been validated by the authors and the manufacturer respectively, to have no cross reactivity with antibodies from other known human coronaviruses.

## Results

There were a total of 7 patients with confirmed COVID-19 admitted to our institution during the study period. A total of 26 samples were retrieved from the Biochemistry department. The number of samples obtained for each patient ranged from 1 – 9 (mean 3.7) with the earliest taken 1 day after the onset of symptoms and latest at day 24. Five specimens were excluded due to the narrow interval between samples or close proximity to the date of onset of illness. Figure 1 summarizes all the test results for these patients. Six out of the 7 patients had at least one positive antibody result and seroconversion was demonstrated in 4 patients. The test results were negative for all the negative control samples except for an IgM IIFT result which was deemed indeterminate due to non-specific fluorescence. An example of a positive result by the IIFA method for IgM and IgG in one of the samples is shown in Figure 2.

**Figure 1:**
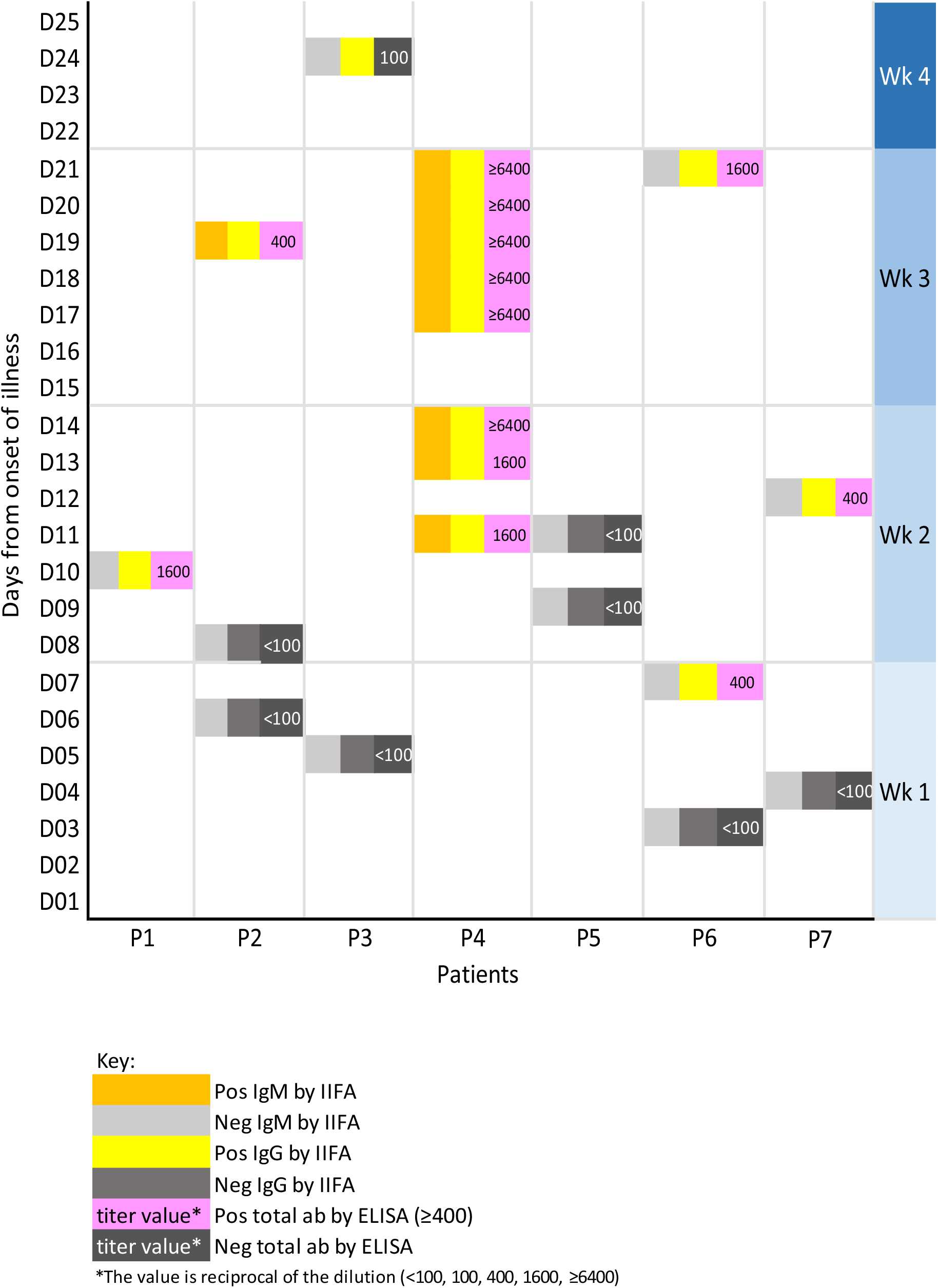
Test results for the COVID-19 patients.

**Figure 2:**
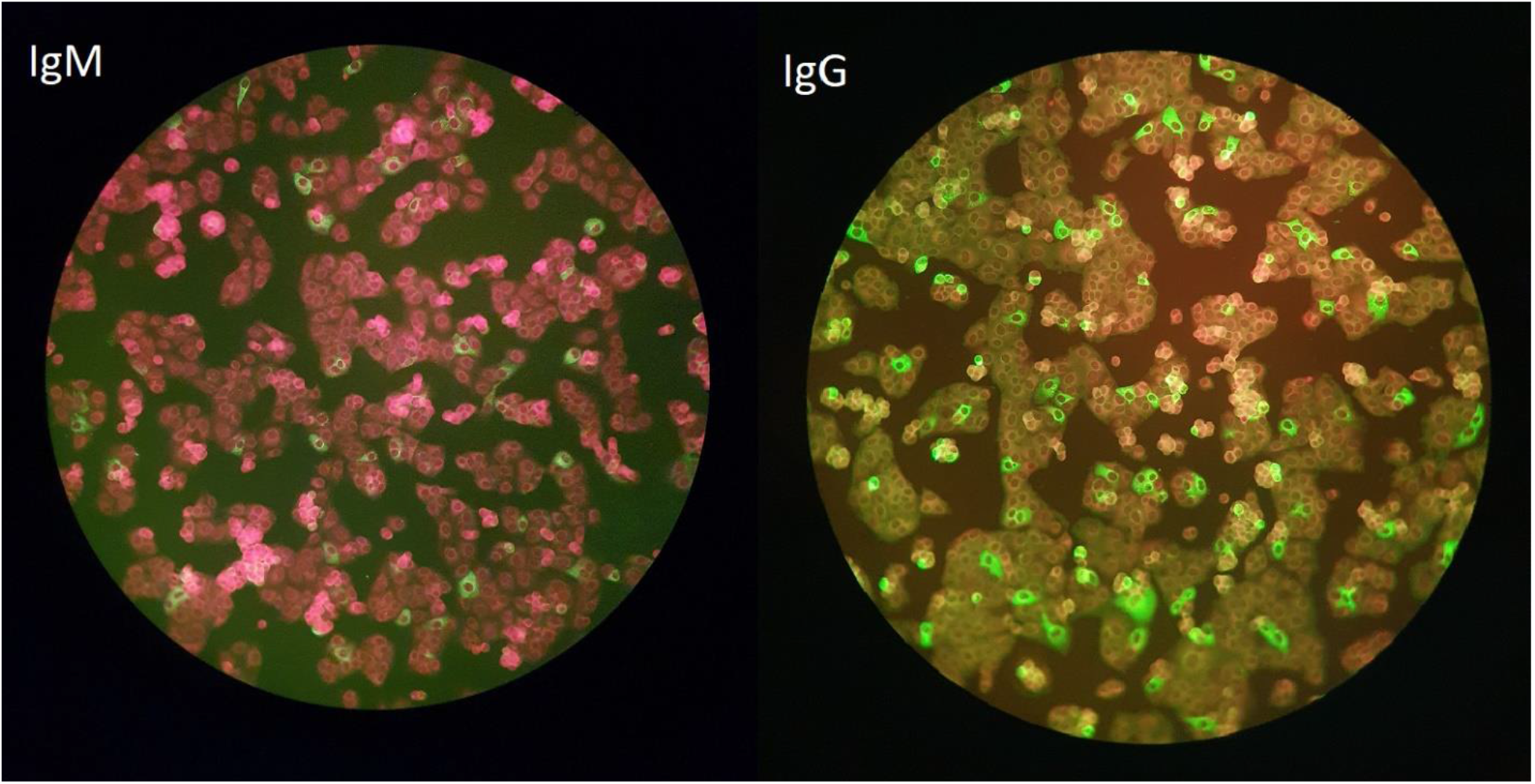
Example of a positive result by the IIFA method for IgM and IgG.

## Discussion

In SARS-CoV, both the IgM and IgG antibodies can appear as early as 1 week after diagnosis in more than half of the patients in one study, with the IgM diminishing from week 5 to undetectable levels by week 11.^9^ Other studies found that 80% of SARS-CoV patients were antibody positive by 8 – 14 days after falling ill and the mean time to seroconversion was 20 days with 93% sensitivity of IgG detection by day 28.^10,11^ Data from our SARS-CoV results of 140 tests by ELISA in 2003 showed that antibodies were positive in 15.4% of samples taken in the 1^st^ week of onset of symptoms, 46.5% in the 2^nd^ week, 88.9% in the 3^rd^ week and a 100% after the 3^rd^ week of onset of illness. For SARS-CoV-2, a study by Zhang et al which used an in- house IgM and IgG ELISA test, found that 50% of their patients were positive for IgM from samples taken on day 0 of hospital admission which increased to 81% by day 5, whereas positive IgG rates increased from 81% to 100%.^12^ However these rates were based on the number of days from the time of hospital admission rather than from the onset of clinical symptoms, hence the early high proportion of positive antibody results reported is not representative of how soon the IgM and IgG appears after infection in this study. In our current COVID-19 cohort (where sera were available for analysis), 25% (1/4) of the patients had detectable antibodies in the 1^st^ week of illness, 66.7% (4/6) by the 2^nd^ week and 100% (5/5) by the 3^rd^ week of illness. These data are somewhat imprecise due to the limited numbers of patients. In addition, we did not have samples from P5 beyond the 2^nd^ week for analysis.

Our evaluation study of both the ELISA and IIFA tests on SARS-CoV patients in 2003 showed that overall the IIFA test was 28.9% more sensitive than the ELISA test, which explains the results of day 24 for P3. Although the overall results for the IIFA test may be more sensitive, the IIFA IgM test in the COVID-19 patients was found to be less useful in the detection of acute phase of the illness which is consistent with the findings of a study in SARS-CoV patients where they found a less frequent (43%) and robust (less discriminatory) IgM response.^13^ However, this cannot be generalized as different assays will have different performances depending on the type of antigen utilized. In most countries, real-time RT-PCR remains the diagnostic tool of choice in the acute phase of infection given that antibody will take time to develop after the onset of illness. These serological tests would be more useful in those who did not present early to a healthcare facility to look for evidence of previous exposure to this virus. This is relevant for contact tracing and to determine the true extent of the circulation of the virus to establish an accurate case-fatality rate.

There is a possibility that positive antibodies from these tests could be as a result of previous exposure to SARS 17 years ago. However, given that only 8,096 cases were reported worldwide and that the virus is not known to still be circulating in the community after it was declared to be contained with no further reported cases in 2004 by WHO, this probability seems very small and can be excluded by specific history taking.

Limitations of this study includes the relatively small number of patients and inconsistent series of sera which ideally should have been collected at a predetermined regular time interval to determine when IgM and IgG can be detected after infection in COVID-19 by these assays. We also did not take into account other factors which could cause the delay in development of antibodies such as immunosuppressive conditions and other treatment modalities that could affect this. However, this study has provided evidence that antibodies to SARS-Cov-2 cross reacts to give positive results in existing SARS-CoV test assays due to the similar structural proteins that it shares with SARS-CoV. The positive predictive value of a serological test depends on the prevalence of the virus and thus in the current situation where there is a recognized outbreak, patients who present with recent compatible symptoms and test positive by these tests are likely to have had exposure to SARS-CoV-2. Serological assays have the advantage in terms of lower set-up costs, capacity for large volume processing, shorter turnaround times, are less prone to specimen sampling quality issues, require lower specific technical skills^14^, have no risk of specimen contamination, involve handling of lower biohazard risk specimen and expose healthcare workers to lower risk during sampling from patients compared to molecular methods.

In conclusion, we provided proof of concept that the available SARS-CoV antibody assays can reliably detect antibodies in patients with COVID-19 which could be used in this current outbreak situation for serosurveys and as a diagnostic tool for under resourced countries. Further studies would be required to confirm their utility and better determine the time frame when IgM and IgG is detectable in patients exposed to SARS-CoV-2.

## Data Availability

Not applicable

## Acknowledgments

We would like to thank the Biochemistry department for their help in retrieving and blinding of specimens, and the Infectious Disease team for providing onset date of illness which was pertinent for the meaningful interpretation of results.

## Notes

### Competing Interest Statement

The authors have declared no competing interest.

### Funding Statement

No external funding was received.

